# Low-Frequency Adaptive Deep Brain Stimulation for Parkinson’s Disease

**DOI:** 10.64898/2025.12.17.25342333

**Authors:** A.S. Negi, C. Cui, K.B. Wilkins, E.F. Lambert, J.A. Melbourne, M.N. Petrucci, S.L. Hoffman, S. Aditham, C. Diep, H.J. Dorris, P. Akella, L. Parisi, A.S. Gala, J.A. Herron, H.M. Bronte-Stewart

## Abstract

**Background:** Freezing of gait (FOG) is a debilitating symptom of Parkinson’s disease (PD) that often worsens with disease progression. Adaptive deep brain stimulation (aDBS) may better target symptom fluctuations and gait impairment, yet most studies have focused on high-frequency stimulation. Low-frequency stimulation may offer additional gait benefits in moderate-to-advanced PD.

**Objective:** To evaluate the efficacy of 60 Hz continuous DBS (cDBS) and the safety, tolerability and feasibility of 60 Hz aDBS.

**Methods:** Eight individuals with PD were implanted with bilateral leads in the subthalamic nucleus and the investigational Summit RC+S system (Medtronic, Inc.). 60 Hz aDBS was calibrated to adjust stimulation amplitude based on prolonged beta bursts. Participants completed blinded, randomized testing during a validated FOG-eliciting stepping-in-place task under OFF DBS, 60 Hz cDBS, and 60 Hz aDBS. Cardinal motor signs were assessed using MDS-UPDRS III in each condition, and under clinical 140 Hz cDBS.

**Results:** 60 Hz cDBS was tolerated acutely by 6 of 8 participants and showed significant shortening of beta burst duration, a pathological neural biomarker of FOG. 60 Hz aDBS was deemed safe and tolerable in this cohort. In participants who demonstrated FOG at baseline, gait improved ON both 60 Hz aDBS and cDBS. However, overall motor symptoms, including tremor, improved only with clinical 140 Hz cDBS.

**Conclusion:** 60 Hz aDBS and cDBS benefit individuals with baseline FOG but may not achieve broad motor symptom relief of clinically optimized HFS. Identifying individuals most likely to benefit from LFS could enable more personalized DBS programming.

## Introduction

Gait impairment and freezing of gait (FOG) are debilitating symptoms of Parkinson’s disease (PD) that increase the risk of falls and impact quality of life and independent living^1,2^. While deep brain stimulation (DBS) of the subthalamic nucleus (STN) generally alleviates motor symptoms in PD, its effect on FOG is often less predictable and may diminish over time as the disease progresses^3,4^. This underscores the need for optimized DBS strategies to specifically address gait impairment and FOG in PD.

Although high-frequency stimulation (HFS, i.e. > 100 Hz) has been clinically established as a stimulation parameter for addressing overall motor symptoms of PD, its effect on gait has been debatable especially for long-term benefit. Low-frequency stimulation (LFS), most commonly at 60 Hz, has been proposed to provide superior benefits on axial motor functions and FOG symptoms over HFS, making it a potential new treatment paradigm for advanced PD with severe gait disorders^5–7^. However, other studies fail to confirm these benefits, showing little-to-no difference between the two frequency settings^8–10^, and some note worsening of symptoms and intolerance to LFS in individuals during acute and long-term evaluation^11,12^. Variability in patient responses suggests that the effects of low-frequency DBS may not be suitable for tremor-dominant individuals^12^. Therefore, the efficacy and feasibility of LFS as a treatment for FOG require further confirmation.

Adaptive DBS (aDBS) therapy has emerged as a promising clinical advancement for managing PD, demonstrating safety, tolerability and efficacy across various studies^13–15^ and now approved by the regulatory bodies in Japan, Europe, and the United States. aDBS automatically modulates stimulation amplitude based on real-time tracking of neurophysiological biomarkers, which are critical in addressing fluctuating symptoms in PD. Previous work showed that freezers exhibited more exaggerated beta band activity in STN than nonfreezers^16,17^, even during non-freezing gait. Furthermore, beta bursts were longer during episodes of FOG compared to during non-freezing walking^16^, indicating that beta band burst duration may serve as a FOG-linked neural biomarker for aDBS. aDBS has predominantly been evaluated using HFS (125-140 Hz). Two studies demonstrated that neural-driven high-frequency aDBS resulted in superior outcomes for sequence effect and FOG compared to cDBS respectively^18,19^. Previous research indicated that low-frequency continuous (c)DBS shortened pathological beta burst durations in the STN and improved FOG^16^. These findings suggest that low-frequency aDBS based on beta burst duration could be a viable alternative and effective strategy to treat gait impairment and FOG in PD. While low-frequency aDBS has been evaluated in a single patient^20^, further data is needed to better evaluate its feasibility and effects on gait symptoms. To fill this gap, this study aimed to investigate the potential benefits of neural driven aDBS at 60 Hz to specifically target gait impairment and FOG.

The aims of the present study were to (i) further assess the efficacy of 60 Hz STN cDBS and (ii) implement and evaluate the feasibility, safety, and efficacy of a beta burst-driven 60 Hz aDBS controller in individuals with moderate-to-advanced PD who experience gait impairment and/or FOG. Quantitative assessments of gait, as well as overall PD motor symptoms, were investigated in this double blinded, randomized study.

## Material and methods

### Participants

Inclusion criteria included a score ≥ 1 on the Freezing of Gait Questionnaire (FOG-Q) or the MDS-UPDRS III gait sub-score (Item 3.10). Participants had bilateral STN DBS leads (model 3389, Medtronic, Inc), and all received the investigational Summit RC+S (IPG, Medtronic, Inc.) either as their first implantable pulse generator (IPG) or when they had an IPG replacement. Full description of the preoperative selection criteria and surgical technique are provided in the Supplementary Material. All participants provided written consent for the study, which was approved by the FDA under an investigational device exemption and by the Stanford University Institutional Review Board.

### Experimental Protocol

All participants were clinically optimized on DBS (140 Hz cDBS) by their neurologist prior to starting research. The research visit was coordinated with a clinical visit in the same week, during which participants were evaluated on their clinical cDBS settings using the MDS-UPDRS Part III. All assessments were conducted in the off-medication state; medication washout protocol is detailed in the Supplementary Material.

### Acute 60 Hz Tolerability Testing

As all participants were clinically treated with 140 Hz cDBS, tolerability to 60 Hz cDBS was first assessed. Stimulation amplitude at 60 Hz was gradually increased from 0 mA to clinical stimulation amplitude. If a patient requested to return to 140 Hz cDBS due to non-transient adverse sensations from the stimulation, they were deemed intolerant to 60 Hz cDBS, and the protocol was suspended. If they tolerated 60 Hz cDBS, they continued with the protocol and were continually monitored for comfort and tolerability.

For participants tolerating 60 Hz cDBS at the clinical amplitude, stimulation amplitude was increased to determine the maximum tolerable stimulation amplitude (i.e. supra-clinical) at 60 Hz. For safety, the amplitude was capped at 125% of 140 Hz cDBS amplitude to avoid potential adverse events.

### Gait Assessment

Quantitative gait measurement used a harnessed stepping-in-place (SIP) task which was previously validated as a measure of FOG^21^. Participants performed 100 seconds of continuous, self-paced alternating stepping within a stationary visual surround that minimized optic flow (Bertec Corporation, Columbus, OH, USA).

### Calibration of aDBS

aDBS programming followed the pipeline previously implemented in a high-frequency beta burst-driven aDBS study^22^. First, participants performed the baseline SIP task during the OFF DBS state. DBS was turned off for at least 15 minutes prior in order to wash out the stimulation effect^23^. Participants then performed the SIP task under randomized stimulation amplitude titrations at 60 Hz cDBS. The stimulation amplitude levels ranged from 50% of clinical amplitude to the participant’s maximum tolerable supra-clinical amplitude.

#### Setting safe and tolerable DBS amplitude limits

A therapeutic window, encompassing a minimum stimulation amplitude (Imin) and a maximum amplitude (Imax), was established based on the participant’s gait performance from baseline and stimulation titrations. Imin was set as a non-zero cDBS amplitude that provided an acceptable therapeutic benefit to the participant’s gait compared to the OFF DBS state. Imax was set as a supra-clinical amplitude that did not result in a deterioration of gait performance; if no such amplitude was identified, Imax was set to the clinical amplitude.

#### Setting safe and tolerable ramp rates

Appropriate ramp rates for adjusting stimulation amplitude within the therapeutic window was established through a random adapting amplitude test^24^. A ramp-up rate of 0.1 mA/s and a ramp-down rate of 0.05 mA/s were initially applied, if not perceptible to participants. If transient side effects occurred (e.g. paresthesias), the rates were decreased until a suitable level was identified, with the ramp-down rate set to half of the determined ramp-up rate to bias stimulation amplitude up and avoid artifactual beta rebound^25,26^.

#### Determining neural input - single threshold beta burst duration

STN local field potentials (LFP) were recorded using the ‘sandwiching’ configuration, with sensing electrodes flanking the stimulating electrode(s), to reduce stimulation-related artifact^27^. LFPs were sampled at 500 Hz. One onboard high-pass filter at 0.85 Hz, one onboard low-pass filter at 100 Hz, and one onboard low-pass filter at 50 Hz were applied^27^. Power spectral analysis was conducted for each STN-LFP to establish the frequency within the beta band (13-30 Hz) at which there was maximum beta band power and exhibited the greatest attenuation during DBS amplitude titrations. A 6 Hz band centered around that frequency was then used to measure beta band burst duration^28^. A single threshold for burst duration was set for each STN to independently drive stimulation^29,30^. The initial burst duration threshold was informed by the mean burst duration during OFF DBS or at the Imin.

#### aDBS implementation

aDBS was implemented using the distributed mode of the Summit RC+S system (Medtronic, Inc.). The computer-in-the-loop system architecture and the custom beta-burst driven control policy algorithm have been previously published^22,30^. The established therapeutic window, ramp rates, and pathological burst duration threshold were implemented into the single-threshold control policy algorithm^22,30^ which modulated stimulation amplitude over time while holding stimulation frequency at 60 Hz and pulse width at 60 µs. Prior to blinded randomized testing, calibration run(s) of aDBS were conducted while the participant performed SIP task, to fine tune burst duration threshold based on observed stimulation temporal dynamics, gait performance, and participant feedback.

### Randomized Blinded Testing

aDBS was evaluated in comparison to cDBS in randomized order. To ensure an effective comparison, cDBS was configured to match the total electrical energy delivered (TEED) by aDBS. Specifically, the cDBS stimulation amplitude was set to the average observed value during the calibration runs of aDBS. The same stimulating electrodes, frequency (60 Hz), and pulse width (60 µs) were used for aDBS and cDBS stimulation conditions.

Each condition was evaluated after at least 20-minute wash-in period. Participants performed the SIP task at the 20-minute timepoints of each stimulation condition and were subsequently assessed with the MDS-UPDRS III by a certified rater. The order of stimulation conditions was blinded to the participant and the MDS-UPDRS III rater.

### Data Acquisition

LFP and stimulation data were streamed off the Summit RC+S device to the computer-in-the-loop via the CTM and structured into JSON files for offline analysis.

Kinetic data was obtained from the dual force plates during the SIP task (Bertec Corporation, Columbus, OH, USA), sampled at 1000 Hz. Kinematics were measured using wearable inertial measurement units (IMUs, APDM, Inc., Portland, OR) placed bilaterally on the shank segments, sampled at 128 Hz.

Neural, kinetic, and kinematic signals were synchronized through a multi-channel data acquisition board (Power1401) with the Spike software (version 2.7, Cambridge Electronic Design, Ltd., Cambridge, England). Detailed synchronization protocol is provided in the Supplementary Material.

### Data Analysis

LFPs and stimulation data were time-aligned using an open MATLAB toolbox^31^. Power spectral density (PSD) estimates for LFPs were calculated using Welch’s method with a 1-s Hanning window and 50% overlap. To calculate beta burst duration, the LFP signal was band-pass filtered within the participant-specific 6 Hz band using a zero-phase, 8th-order filter, and the output was squared to create a power envelope. The power threshold for detecting beta bursts was determined using the median power of the troughs of a gamma band (45 to 65 Hz) envelope, and was set at four times that median^28^. Burst durations were then calculated as the time interval between successive crossings of this threshold by the beta power envelope.

Vertical ground reaction forces under each foot were low-pass filtered with a 20 Hz cut-off frequency and normalized to individual body weight. FOG episodes during SIP were identified using a validated custom algorithm^21^. Tri-axial gyroscope data from IMU was low-pass filtered with a 9 Hz cut-off frequency. Principal component analysis was used to extract the 1-D shank angular velocity within the sagittal plane. Shank swings were identified by detecting shank angular velocity peaks above the threshold of 10 deg/s. Gait arrhythmicity was calculated as the natural log transformation of stride time coefficient of variation (CV).

### Statistical analysis

Statistical analyses were run in R (version 3.6.0, R Foundation for Statistical Computing, University of Auckland, New Zealand). A linear mixed effects model with a fixed effect of titration level and random effects of participant and STN side were conducted to assess the effect of stimulation on mean burst duration. Linear mixed effects models with a fixed effect of stimulation condition and random effect of participant were run for each of the motor outcomes, including the MDS-UPDRS III scores (total score, tremor, bradykinesia, and rigidity sub-score), and SIP task metrics (percent time freezing, arrhythmicity, and peak shank angular velocity). Effect size was reported with Partial eta squared (ηp^2^). Estimated marginal means were computed for post hoc pairwise comparisons between stimulation conditions with Tukey’s adjustment. Significance was set at *p* < 0.05.

## Results

### Participant Demographics

Eight participants were enrolled. Their demographics and preoperative characteristics are listed in Table 1. One participant was a non-freezer with mild gait impairment as evident by a score of 1 on the MDS-UPDRS III gait item 3.10; the other seven participants were freezers based on FOG-Q or New FOG-Q.

**Table 1.** Demographics.

| ID | Sex | Age (years) | Disease Duration (years) | FOG-Q (*)/<br>New FOG-Q | MDS-UPDRS III<br>Gait | IPG | HFS Duration (years) |
| --- | --- | --- | --- | --- | --- | --- | --- |
| P1 | M | 60-64 | 10 | 0* | 1 | 1<sup>st</sup> | 1.3 |
| P2 | M | 60-64 | 15 | 2* | 2 | 3<sup>rd</sup> | 7.9 |
| P3 | F | 70-74 | 16 | 4* | 3 | 1<sup>st</sup> | 1.8 |
| P4 | M | 65-59 | 13 | 2* | 2 | 1<sup>st</sup> | 1.5 |
| P5 | M | 65-69 | 11 | 3* | 1 | 3<sup>rd</sup> | 7.8 |
| P6 | F | 60-64 | 20 | 14 | 2 | 3<sup>rd</sup> | 6.6 |
| P7 | M | 70-74 | 16 | 25 | 1 | 1<sup>st</sup> | 1.5 |
| P8 | F | 55-59 | 14 | 15 | 1 | 3<sup>rd</sup> | 7.1 |

FOG-Q = Freezing of Gait Questionnaire; NFOG-Q = New Freezing of Gait Questionnaire; IPG = implantable pulse generator; HFS = high-frequency stimulation.

Figure 1 shows the lead location for all participants. Four participants received the Summit RC+S as their first IPG, and four others as a replacement IPG. Participants had been clinically treated with 140 Hz stimulation, with durations ranging from 1.3 to 7.9 years (Table 1).

**Figure 1.**
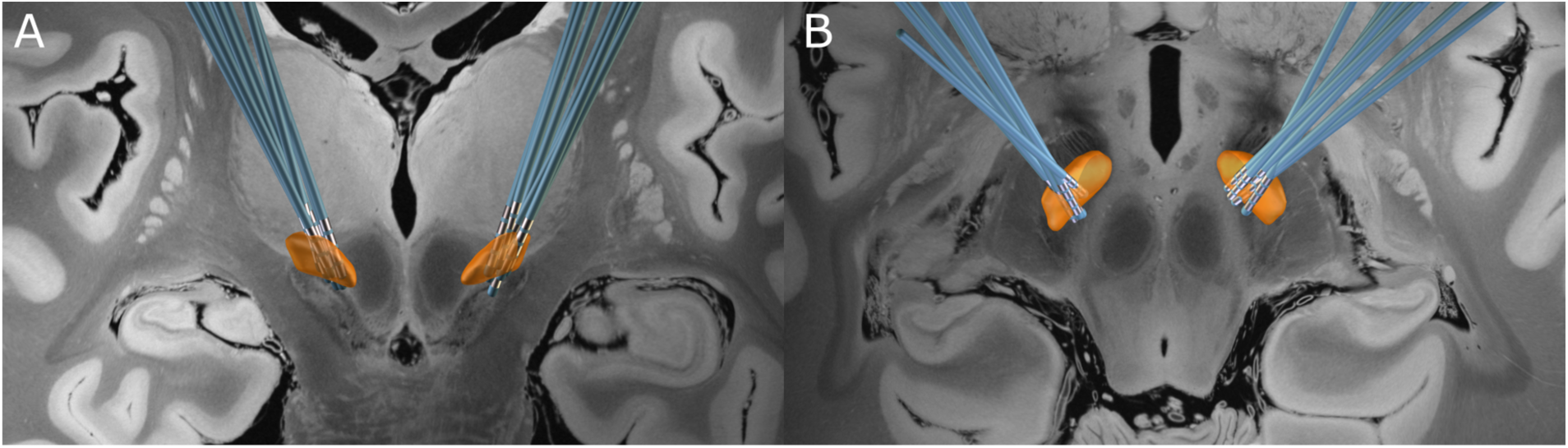
Lead locations. Locations of bilateral leads in STN for all eight participants. (A) Coronal and (B) Axial views.

### Tolerability to 60 Hz

Six of the eight participants were able to tolerate 60 Hz cDBS acutely, while two other participants experienced immediate return of PD symptoms, preventing further testing. Of the participants who were unable to tolerate 60 Hz acutely, one experienced chest stiffness (P2) and another reported return of tremor and stiffness (P8). One participant (P6) tolerated 60 Hz acutely but experienced worsening of PD symptoms and intense discomfort over longer exposure to 60 Hz settings during experiments; however, they were able to complete the gait and MDS-UPDRS III testing.

### 60 Hz aDBS Calibration

The six participants who could tolerate 60 Hz cDBS acutely were set up with LFP sensing and aDBS programming. Four participants were configured with bilateral LFP sensing, while two other participants were configured with unilateral sensing (Table 2). Participant 4’s RSTN signals had stimulation-induced artifacts which prevented obtaining reliable neural metrics; Participant 5’s RSTN had the ventral contact as stimulating contact, and therefore a sandwiching sensing configuration was not possible. As a result, the two STNs were maintained at continuous stimulation at clinical amplitude during the aDBS condition.

**Table 2.** Individual 60 Hz aDBS and cDBS parameters.

| ID | Center Frequency (Hz) |  | Burst Duration Threshold (ms) |  | Therapeutic Window (mA) |  | Ramp Rates Up/Down (mA/s) |  | cDBS Stim Amplitude (mA) |  |
| --- | --- | --- | --- | --- | --- | --- | --- | --- | --- | --- |
|  | LSTN | RSTN | LSTN | RSTN | LSTN | RSTN | LSTN | RSTN | LSTN | RSTN |
| P1 | 24.4 | 17.6 | 90 | 110 | 1.8-2.4 | 1.8-2.4 | 0.1/0.05 | 0.1/0.05 | 2.3 | 2.0 |
| P3 | 13.7 | 12.7 | 310 | 142 | 1.3-3.1 | 1.4-3.4 | 0.06/0.03 | 0.06/0.03 | 2.8 | 3.4 |
| P4 | 17.6 | - | 150 | - | 2.4-3.2 | - | 0.1/0.05 | - | 3.1 | 4.2 |
| P5 | 16.6 | - | 450 | - | 5.4-5.9 | - | 0.08/0.04 | - | 5.7 | 6.7 |
| P6 | 25.4 | 25.4 | 250 | 200 | 4.2-6.0 | 4.2-6.0 | 0.1/0.05 | 0.1/0.05 | 5.8 | 5.9 |
| P7 | 16.6 | 15.4 | 700 | 200 | 1.9-2.9 | 1.3-2.1 | 0.1/0.05 | 0.1/0.05 | 2.7 | 1.9 |

Dash indicates that the STN was held at cDBS at clinical amplitude

The center frequencies for the individual beta bands are shown in Table 2. Analysis of beta burst duration across OFF and stimulation titration levels revealed a statistically significant effect of stimulation amplitude on burst duration where increased levels of 60 Hz stimulation amplitude led to decreased beta burst durations (β = −0.005, CI= [−0.007 −0.003], *p* < 0.001, ηp^2^ = 0.19; Figure 2).

**Figure 2.**
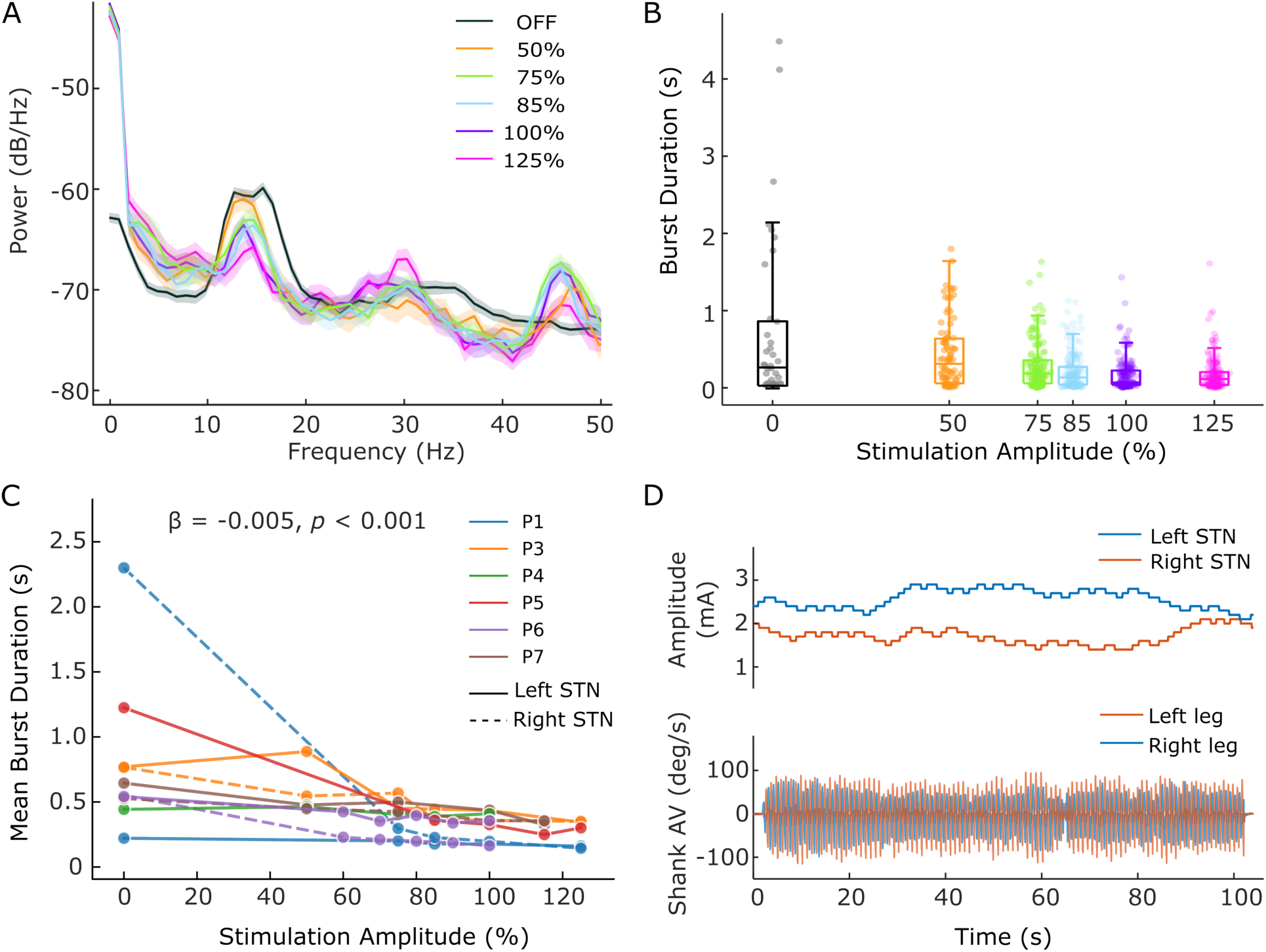
STN-LFP analysis and aDBS implementation. (A) Example power spectral density of local field potential during gait OFF stimulation and ON various titration levels. Stimulation amplitude is expressed as a percentage of clinical amplitude. A 6-Hz band centered around the frequency which exhibited the most prominent modulation with stimulation titrations (in this example 13.7 Hz) was selected for tracking beta bursts. (B) Boxplots of duration of beta bursts at each stimulation amplitude level. (C) Mean beta-burst duration for each participant across the stimulation titration levels. (D) Example aDBS implementation during gait, showing synchronized recordings of stimulation amplitude and shank angular velocity measured from IMU sensors.

60 Hz aDBS was successfully implemented across the six participants who tolerated 60 Hz cDBS. Four participants tolerated the ramp-up rate of 0.1 mA/sec and ramp-down rate of 0.05 mA/sec with no sensation of ramping, whereas two reported adverse sensations at 0.1 mA/s and therefore received slightly slower ramp rates (Table 2). Fig. 2D depicts the temporal dynamics of an example aDBS run with synchronous recording of stimulation amplitude and gait kinematics (see Supplementary Video). 60 Hz aDBS was safe and well tolerated in all six individuals.

### 60 Hz cDBS and aDBS on Gait and FOG

Table 2 listed individual stimulation amplitude for 60 Hz cDBS condition, which was configured to match the TEED of 60 Hz aDBS.

Randomized blinded comparison revealed variable responses to 60 Hz aDBS and 60 Hz cDBS compared to OFF DBS across participants (Figure 3). Three freezers (P3, P4, P5) displayed freezing during SIP OFF DBS, while the non-freezer (P1) and two freezers (P6, P7) did not freeze. All three participants who froze OFF DBS showed improvement ON both 60 Hz cDBS and aDBS: reduced percent time freezing, reduced gait arrhythmicity and increased shank angular velocity.

**Figure 3.**
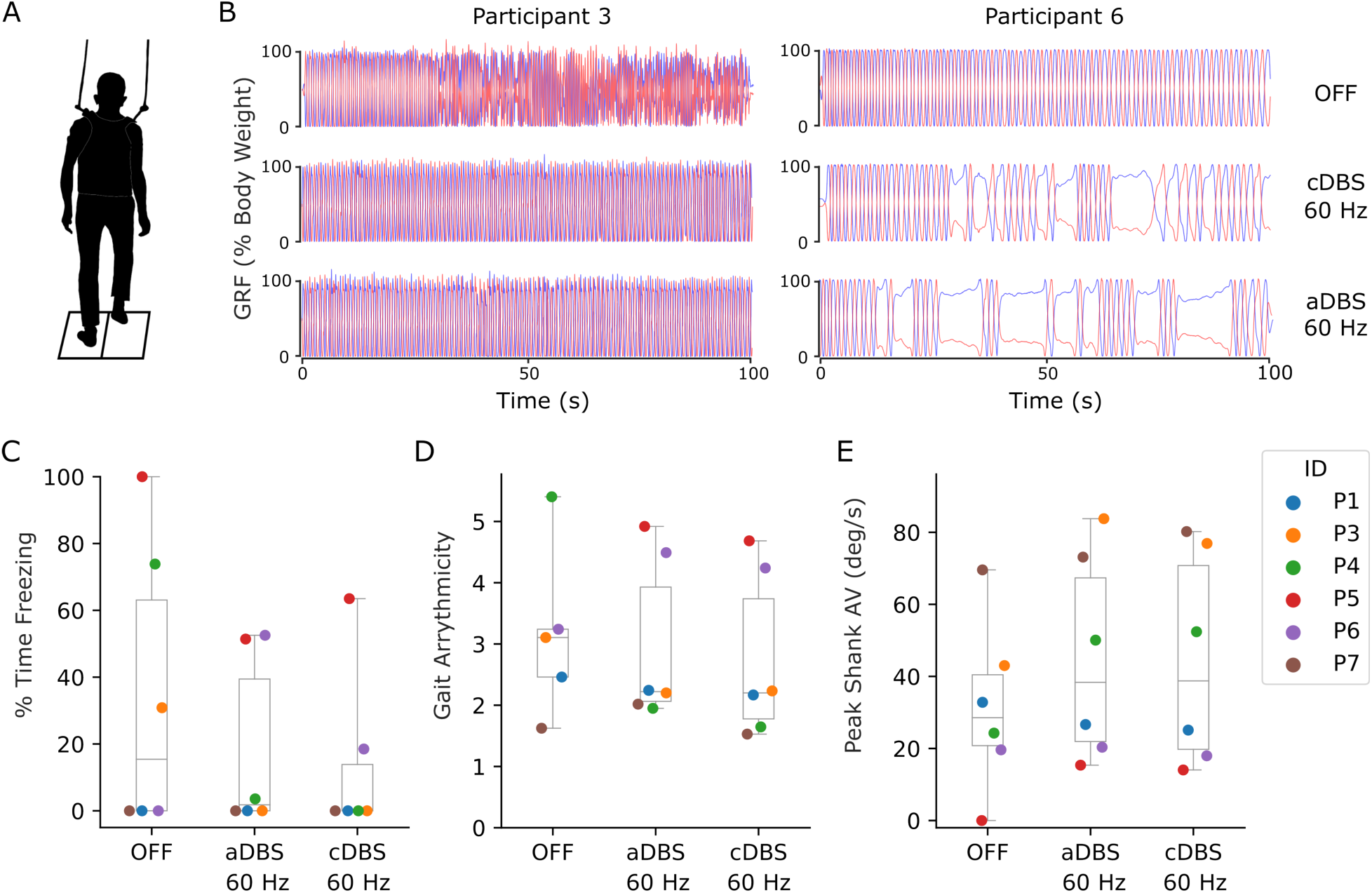
Quantitative gait measures. (A) Depiction of the SIP task. (B-C) Example SIP task performance for a responder (P3) and a non-responder (P6) to 60 Hz stimulation conditions. Vertical ground reaction forces (GRFs) are shown as a percentage of body weight. Nonfreezing stepping is indicated by continuous full weight shifting between limbs. (B) Participant 3 displayed freezing behavior during OFF DBS and no freezing while ON 60 Hz DBS. (C) Participant 6 did not freeze during OFF DBS but exhibited long episodes of freezing ON 60 Hz DBS. (D-F) SIP task metrics for all six participants across OFF DBS, 60 Hz aDBS, and 60 Hz cDBS, including percent time freezing, gait arrhythmicity, and peak shank swing angular velocity. Colors represent individual participants. Participant 5 could not ambulate OFF DBS, therefore no gait arrhythmicity (E) obtained for that trial.

Of the three participants who did not freeze during SIP OFF DBS, the non-freezer and one freezer also did not freeze ON 60 Hz cDBS or aDBS, whereas one freezer (P6) froze ON both 60 Hz cDBS and aDBS, and demonstrated increased arrhythmicity compared to OFF DBS.

On the group level, no statistically significant differences were found across the three conditions (percent time freezing: F_2,8.8_ = 1.45, *p* = 0.286, ηp^2^ = 0.25; arrhythmicity: F_2,8.1_= 0.35, *p* = 0.712, ηp^2^ = 0.08; peak shank angular velocity: F_2,10_= 3.47, *p* = 0.072, ηp^2^ = 0.41).

### Overall Clinical Motor Signs

Figure 4 presents the overall motor symptom evaluation using MDS-UPDRS III during the randomized blinded testing, along with a clinical 140 Hz cDBS evaluated during the clinic visit.

**Figure 4.**
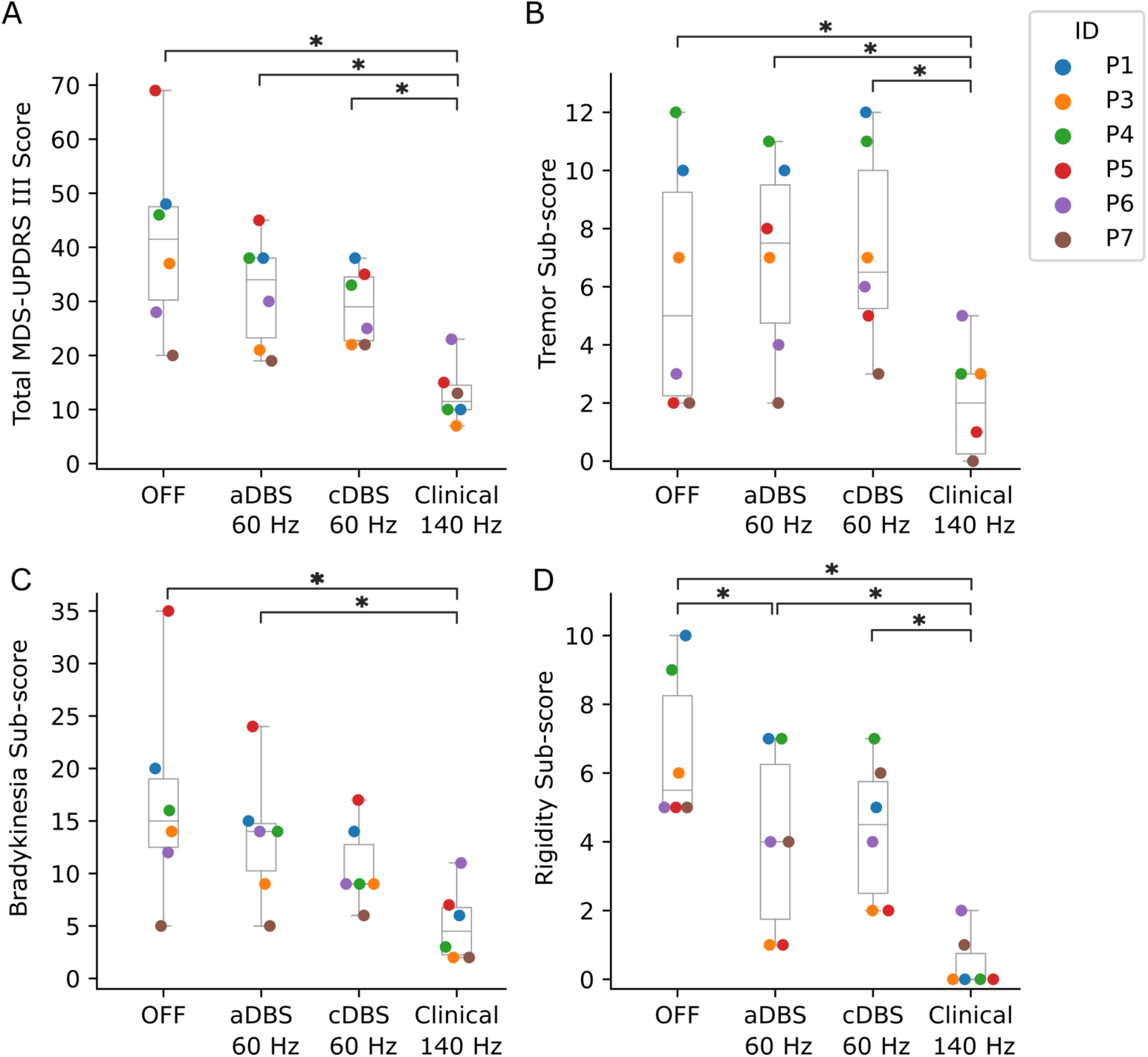
MDS-UPDRS III evaluations. (A) Total score and (B) Tremor (C) Bradykinesia (D) Rigidity sub-scores across OFF stimulation, 60 Hz aDBS, 60 Hz cDBS, and patient’s clinically optimized cDBS at 140 Hz. Colors represent individual participants. Asterisks denote the significant difference between conditions according to the post hoc pairwise comparisons.

A significant main effect of stimulation condition was observed for the MDS-UPDRS III total score (F_3,15_ = 11.7, *p* < 0.001, ηp^2^ = 0.70). Post hoc pairwise comparisons showed that 60 Hz aDBS and 60 Hz cDBS did not show improvement compared to OFF DBS. In contrast, ON their clinical 140 Hz cDBS settings, participants’ total scores were significantly better compared to OFF DBS (*p* < 0.001) and compared to ON both 60 Hz conditions (aDBS: *p* = 0.007, cDBS: *p* = 0.021).

A significant main effect of stimulation conditions was observed for MDS-UPDRS III tremor (F_3,15_ = 6.5, p = 0.005, ηp^2^ = 0.56), bradykinesia (F_3,15_ = 8.5, *p* = 0.002, ηp^2^ = 0.63), and rigidity sub-scores (F_3,15_ = 15.8, p < 0.001, ηp² = 0.76). For tremor, 60 Hz aDBS and cDBS did not show improvement compared to OFF DBS, while clinical 140 Hz showed significant improvement compared to OFF (*p* = 0.046) and 60 Hz DBS conditions (aDBS: *p* = 0.011, cDBS: *p* = 0.007). For bradykinesia, 60 Hz DBS conditions did not show improvement compared to OFF DBS. Bradykinesia improved ON clinical 140 Hz DBS relative to OFF DBS (*p* = 0.001) and to 60 Hz aDBS (*p* = 0.017) but not to 60 Hz cDBS (*p* = 0.150). Rigidity showed improvement ON 60 Hz aDBS compared to OFF (*p* = 0.044) and ON clinical 140 Hz compared to OFF (*p* < 0.001) and to both 60 Hz DBS conditions (aDBS: *p* = 0.007, cDBS: *p* = 0.004).

## Discussion

This study provides the first investigation of 60 Hz STN aDBS in a PD cohort with gait impairment and FOG and demonstrated that beta burst-driven aDBS at 60 Hz was safe, feasible and tolerable among individuals who could tolerate 60 Hz cDBS. 60 Hz aDBS and cDBS effectively shortened beta burst duration and improved FOG in the participants who demonstrated FOG OFF DBS.

The non-freezer (P1) in the study had normal gait arrhythmicity and shank angular velocity OFF DBS, and these did not change ON 60 Hz aDBS or cDBS. However, two participants, who were classified as freezers, did not freeze during the baseline SIP OFF DBS. Of these, one also did not freeze ON 60 Hz aDBS and 60 Hz cDBS, but the other participant experienced increased FOG and arrhythmicity ON both 60 Hz DBS conditions. This does not necessarily imply that 60 Hz DBS caused FOG as this participant was identified as a freezer and only had one SIP test performed OFF DBS.

The differential effects of 60 Hz on gait, seemingly associated with the presence or absence of baseline FOG, suggest that its therapeutic benefit of 60 Hz may be specific to FOG. These observations are consistent with our previous study^16^ which showed that both 60 Hz and 140 Hz cDBS shortened pathological beta burst durations in freezers and improved FOG, but left unchanged the beta burst durations and normal gait parameters in non-freezers. Together, the results suggest that 60 Hz’s benefit may be specific to FOG-linked pathological beta activity.

One well-documented concern regarding LFS for PD is that it tends to be ineffective in managing tremor^8,12^. This study demonstrated that 60 Hz DBS provided no therapeutic benefit for tremor and even worsened symptoms in some participants compared to OFF, whereas clinical 140 Hz cDBS improved tremor as well as the other cardinal motor symptoms. Other studies have also documented intolerance to LFS in some individuals, citing tremor worsening as the key reason^12,32^. The lack of benefit and potential worsening of tremor ON LFS may preclude tremor-dominant individuals with FOG from receiving clinical benefit from 60 Hz aDBS or cDBS.

The MDS-UPDRS III total score and bradykinesia subscore also did not improve ON 60 Hz aDBS or 60 Hz cDBS compared to OFF DBS. Rigidity improved only ON 60 Hz aDBS compared to OFF DBS. In contrast, overall MDS-UPDRS III, bradykinesia, tremor, and rigidity all improved ON clinical 140 Hz cDBS compared to OFF DBS and compared to 60 Hz aDBS. All but bradykinesia showed significant improvement on clinical 140 Hz cDBS compared to 60 Hz cDBS. These results highlight that while 60 Hz may have efficacy on FOG, cardinal motor symptoms were better treated ON 140 Hz cDBS. One possible explanation is that HFS effectively suppresses broad beta oscillations which are linked to motor dysfunction^33,34^. In contrast, 60 Hz DBS has been shown to attenuate high-beta (19–27 Hz) but amplify alpha/low-beta activity (11–15 Hz)^35,36^, which may reflect effective decoupling of hyper-direct STN-motor cortices pathways but increased coupling in the cortico-striato-basal ganglia network. Such differential modulation could underlie why 60 Hz stimulation significantly shortened beta burst at the patient-specific beta frequency (6 Hz band) yet lacked control of or even worsened cardinal motor symptoms and tremor. Further research is warranted to characterize the specific frequency range and degree of modulation by LFS versus HFS, particularly in individuals with long disease duration and/or exposure to DBS.

Adverse responses to 60 Hz stimulation presented heterogeneously across the three individuals (P2, P6, P8), including increased rigidity, tremor, and worsening in gait and axial symptoms. Examining the demographics, all three had been on clinical HFS for a longer duration (7.2 ± .65 years) with a disease duration of 16 ± 3.2 years, compared to those who could tolerate 60 Hz (2.8 ± 2.8 years on HFS and 13.2 ± 2.8 years with PD) (Table 1). This suggests that longer duration on HFS may be a predictor of intolerance and/or symptom worsening to LFS. This is supported by previous evidence^37,38^ where LFS improved patient outcomes when initiated early in STN-DBS programming (i.e. within a few months post surgery). A similar trend has been documented for pallidal-DBS for dystonia, where a lack of benefit from pallidal 60 Hz DBS was associated with longer duration of symptoms, exposure to HFS as well as older age at the time of surgery^39,40^. The mechanisms underlying potential development of intolerance to LFS following prolonged HFS are interesting and warrants further exploration. Alternatively, LFS may offer the most therapeutic benefit when it lessens the detrimental effects of HFS in select patients^41^, and the present cohort already demonstrated favorable responses to HFS (Fig. 4), which may explain the limited efficacy of 60 Hz stimulation in this group. Future studies could explore the efficacy of low-frequency aDBS in select patient cohorts, potentially with short exposure to HFS and/or suboptimal response to HFS.

### Limitations

First, this study had a small sample size due to an unexpected discontinuation of the investigative Summit RC+S in October 2022. Nevertheless, it represents the largest PD cohort studied to date with 60 Hz aDBS for gait impairment and FOG. Second, because aDBS requires a ‘sandwich’ configuration for LFP sensing, clinical equivalent DBS settings were used in six STNs across four participants with modified stimulation contact and amplitude (Supplementary Table S1). Third, the protocol lacked a 140 Hz cDBS condition in the randomized quantitative testing. The condition was omitted in protocol design to minimize fatigue. Instead, MDS-UPDRS III ratings under 140 Hz cDBS from the clinic visit were incorporated as a benchmark for comparing overall motor symptoms. Lastly, the stimulation amplitude at 60 Hz was capped at 125% of patient’s clinical amplitude for safety purposes, which corresponded to a lower TEED compared with clinical 140 Hz cDBS. However, previous studies documented positive responses to LFS without changes in stimulation amplitude, contact or pulse width^7,37,42^, indicating the therapeutic effect was associated with the frequency regardless of TEED.

## Conclusion

This is the first quantitative, objective investigation of low-frequency STN aDBS in a cohort of PD individuals with gait impairment and FOG. 60 Hz aDBS was safe and tolerable in patients who tolerate 60 Hz cDBS. 60 Hz aDBS improved FOG and showed similar efficacy to 60 Hz cDBS. However, overall MDS-UPDRS III scores were worse on 60 Hz cDBS and aDBS compared to clinical 140 Hz DBS and showed no improvement compared to OFF DBS. Individuals who could not tolerate 60 Hz cDBS tended to have had longer exposures to 140 Hz cDBS. These findings establish the feasibility of 60 Hz aDBS for PD+FOG and pave the way for future research into low-frequency aDBS in individuals who are most likely to benefit from LFS. Of note, 60 Hz DBS may not be a viable therapy option for advanced PD patients who have undergone prolonged exposure to HFS and/or who present with a range of cardinal motor symptoms, including tremor.

## Data Availability

All data produced in the present study are available upon reasonable request to the authors

## Acknowledgements

The authors greatly appreciate the participants who dedicated their time to this study. We also thank the Open Mind Consortium for providing technical support in processing Json data from the Summit RC+S. This work was funded by NINDS UH3NS107709, UH3NS128150, U24NS113637, Robert and Ruth Halperin Foundation, John A. Blume Foundation, John E. Cahill Family Foundation, and Medtronic Inc. who provided devices but no additional financial support.

