## Supplementary Material for "Low-Frequency Adaptive Deep Brain Stimulation for Parkinson’s Disease"

Supplementary Methods

*Pre-op inclusion and exclusion criteria*

Preoperative inclusion criteria are being at least 18 years of age, meeting criteria for STN DBS, presence of complications of medication such as wearing off signs, fluctuating responses, dyskinesias, medication refractory tremor and/or impairment in the quality of life on or off medication, and a score ≥ 1 on the freezing of gait questionnaire (FOG-Q) and/or gait sub-score (Item 3.10) of Movement Disorders Society Unified Parkinson’s Disease Rating Scale Part III (MDS-UPDRS III) in the off or on state. Exclusion criteria included being over the age of 80, dementia, untreated psychiatric disease, Hoehn and Yahr stage 5, major surgical morbidities, presence of a cardiac pacemaker, required rTMS, ECT, MRI, or diathermy, pregnancy, cranial metallic implant or history of seizures or epilepsy.

*Surgical procedure*

PD subjects underwent bilateral implantation of DBS leads (model 3389, Medtronic, Inc.) in the sensorimotor region of the STN using standard frameless stereotactic technique, with multi-pass microelectrode recording (MER)^1^, and test neurostimulation through the cannula around the microelectrode as well as through the DBS lead. Leads were connected to the investigative neurostimulator Summit™ RC + S (Medtronic, Inc.) which was implanted in the right chest.

*Electrode localization*

Preoperative T_1_ and T_2_ MRI scans and postoperative CT scans were acquired as part of the standard Stanford clinical protocol^1^. Location of DBS leads was determined by the Lead-DBS toolbox^2^. Postoperative CT scans and preoperative T_2_ scans were co-registered to preoperative T_1_ scans, which were then normalized into MNI space using SPM12 (Statistical Parametric Mapping 12; Wellcome Trust Centre for Neuroimaging, UCL, London, UK) and Advanced Normalization Tools^3^. DBS electrode localizations were then corrected for brain-shift in the postoperative CT scan^4^. DBS electrodes were then localized in template space using the PaCER algorithm^5^ and projected onto the DISTAL Atlas to visualize overlap with the STN^6^.

*Medication washout protocol*

Both clinical and research visits were conducted with participants in the off-medication state, which involved discontinuing long-acting dopamine agonists for at least 48 hours, dopamine agonists and controlled-release carbidopa/levodopa for at least 24 hours, and short-acting medication for at least 12 hours.

*Multi-model data synchronization*

Neural, kinetic, and kinematic signals were synchronized using the Spike software (version 2.7, Cambridge Electronic Design, Ltd., Cambridge, England) through a multi-channel data acquisition board (Power1401). The IMU system automatically sent a TTL pulse to the data acquisition board at the start of recording. For synchronizing force plate and LFP data, we connected external instruments to the data acquisition board. Force plate data were synchronized using a rapid hammer strike, which was recorded simultaneously by both the force plate and an accelerometer. LFP data were synchronized by delivering a 20 Hz stimulation pulse train, which was recorded by the IPG and a surface electrode placed over the patient IPG wire.

Supplementary Results

**Table S1**. Individual stimulation configuration for clinical DBS and modified aDBS contacts, clinical DBS and clinical equivalent DBS amplitude, and cDBS amplitude.

| ID | Clinical Contacts | Clinical Amplitude | Clinical Equivalent Contacts | Clinical Equivalent Amplitude | 60 Hz cDBS  Amplitude |
| --- | --- | --- | --- | --- | --- |
| P1 | LSTN: 1, 2  RSTN: 9 | LSTN: 2.1  RSTN: 2.4 | LSTN: 1  - | LSTN: 2.4  - | LSTN: 2.3 RSTN:2.0 |
| P3 | LSTN: 1, 2  RSTN: 9 | LSTN: 1.25  RSTN:2.7 | LSTN: 2  - | LSTN: 2.6  - | LSTN: 2.8 RSTN:3.4 |
| P4 | LSTN: 1, 2  RSTN: 9, 10 | LSTN: 1.45  RSTN: 2.3 | LSTN: 1  RSTN: 10 | LSTN: 3.2 RSTN:4.2 | LSTN: 3.1 RSTN:4.2 |
| P5 | LSTN: 1, 2  RSTN: 8, 9 | LSTN: 5.8  RSTN:6.7 | -  - | -  - | LSTN: 5.7 RSTN:6.7 |
| P6 | LSTN: 9, 10  RSTN: 0, 1 | LSTN: 3.1  RSTN: 2.65 | LSTN: 9  RSTN: 1 | LSTN: 6.0 RSTN:6.0 | LSTN: 5.8 RSTN:5.9 |
| P7 | LSTN: 2  RSTN: 9 | LSTN: 2.5 RSTN:1.8 | -  - | -  - | LSTN: 2.7 RSTN:1.9 |

Note: All participants had (single/double) monopolar settings. Clinical equivalent settings at 140 Hz were established based on similar efficacy with the patient’s clinical settings. Dash indicates no change from clinical contact/amplitude. cDBS = continuous Deep Brain Stimulation, the 60 Hz experiment condition during the randomized blinded testing, where stimulation amplitudes were modified from their clinical amplitude in order to match the TEED with aDBS condition.

**Table S2**. Individual 60 Hz cDBS and Imax tolerability results

| ID | Feedback | LSTN Clinical | LSTN Imax | RSTN Clinical | RSTN Imax |
| --- | --- | --- | --- | --- | --- |
| P1 | Tolerable | 2.4 | 3.0 | 2.4 | 3.0 |
| P2 | Intolerable | 5.0 | - | 6.6 | - |
| P3 | Tolerable | 2.5 | 3.1 | 2.7 | 3.4 |
| P4* | Tolerable | 3.2 | 3.2 | 4.2 | 4.2 |
| P5 | Tolerable | 5.4 | 6.7 | 5.8 | 7.2 |
| P6 | Tolerable | 6.0 | 6.0 | 6.0 | 6.0 |
| P7 | Tolerable | 2.5 | 2.9 | 1.8 | 2.1 |
| P8 | Intolerable | 3.4 | - | 6.2 | - |

Note: * marks the participant whose Imax was maintained at their clinical HFS amplitude due to adverse sensations at higher amplitudes on 60 Hz DBS
